# How reacted USA with the first case of Bubonic Plague on 21 August?

**DOI:** 10.1101/2021.02.11.21251588

**Authors:** Josimar Edinson Chire Saire, Jimy Oblitas Cruz

## Abstract

The present work studies the reactions of citizens from United States of America (USA) during the first cases of Bubonic Plague in the territory on August 2020, during pandemic generated by Covid-19. The interest of the study is analyze the posts of users from all the states of USA following a Text Mining approach. The collection of data is performed through Twitter Application Program Interface (API) of Twitter, considering keywords: bubonic plague and black death. The results show the states with highest number of coronavirus has more publication than others and the interest about treatments, political issues and public health.

## I. Introduction

In August 2020, a news story is published that a resident of California, in the United States, tested positive for black death, being the first human case of the disease in this American state in five years [1]. Along with this, in December 2019, a new coronavirus triggered a group of cases of severe pneumonia in Wuhan, China. Since then, the World Health Organization (WHO) has called the disease “coronavirus disease 2019” (COVID-19) and the etiological agent “severe acute respiratory syndrome-coronavirus-2” (SARS-CoV-2) [2].

Bubonic plague ranks as the most severe bacterial disease known to man as judged by historical records of mortality and current understanding of the pathological processes that promote the infection [3].

Both diseases are generated by zoonotic agents, so that the population begins to relate both diseases. This case raises a question of how quickly social networks can identify potential outbreaks of zoonotic diseases, especially when comparing with the emergence of COVID-19. The plague identified in USA is a deadly infectious zoonotic disease and is caused by Yersinia pestis, a Gram-negative, immobile cocobacillus, which mainly affects wild rodents, which are its natural reservoirs and can be transmitted to humans and other animals through the bite of fleas from infected rodents. Therefore, it is also known as vector-borne disease [4].

Another important point is to determine the rapid onset of potential risks to public health, such as infections by pathogenic micro-organisms and, because plague in humans is also transmitted by chance contact with infected animal tissue or by the entry of aerosol bacteria by inhalation [5], it is interesting to see how information is disseminated through social networks and to show that this determination can be a useful tool for the immediate detection of risks to public health.

In order to help public health and to make better decisions regarding Public Health and to help with their monitoring, Twitter has demonstrated to be an important information source related to health on the Internet [6], due to the volume of information shared by citizens and official sources. Twitter provides researchers an information source on public health, in real time and globally. Thus, it could be very important for public health research

In this new world scenario, as established by Espina [7], it is necessary to find the determinants of disease outbreaks before they occur, to reduce their impact on populations, and one of the great advantages is to obtain information brought by automated systems.

Therefore, concepts, such as generating a relationship between consumer health informatics and public health informatics, must be taken into account. Currently, info-metrics and web analysis tools are being used for this type of purpose, obtaining information in an electronic means, specifically the Internet, with the ultimate goal of having a positive impact on public health and public policies [8]

## II. Data and Methods

The present work performs experiments with source data from Twitter with Natural Language Processing and Data Mining (Text Mining).

- Choose terms to search on Twitter
- Setup parameters of the query for Twitter and collect data
- Pre-processing data to eliminate words with no relevance (stopwords)
- Visualization

### A. Gather Relevant Terms

After an exploratory of trends and keywords related to Bubonic Plague, the next terms are chosen:

- ‘bubonic plague’,’black death’

,’#bubonicplague’,’#blackdeath’

### B. Setup Parameters for the Query and Collect Data

The extraction of tweets is through Twitter API, with the next parameters:

- date: 14-08-2020 to 21-08-2020
- terms: the chosen words mentioned in previous subsection
- geolocalization, radius: values for each state, see Table. I and Figure 1 for general overview
- language: English

**TABLE I.** Geolocalization of States

| Name | latitude | longitude | radius |
| --- | --- | --- | --- |
| NEVADA | 39.67 | -116.56 | 194.977,11 |
| NEW MEXICO | 33.65 | -106.09 | 234.513,02 |
| NEW YORK | 42.89 | -72.87 | 281.521,85 |
| NORTH CAROLINA | 35.10 | -78.87 | 178.427,53 |
| NORTH DAKOTA | 47.33 | -100.82 | 166.341,38 |
| OHIO | 40.94 | -82.33 | 217.952,92 |
| OKLAHOMA | 35.44 | -97.41 | 174.709,95 |
| OREGON | 42.93 | -121.19 | 234.513,02 |
| SAN FRANCISCO | 38.14 | -121.51 | 252.312,33 |
| SOUTH CAROLINA | 32.84 | -80.86 | 123.181,40 |

**Fig. 1.**
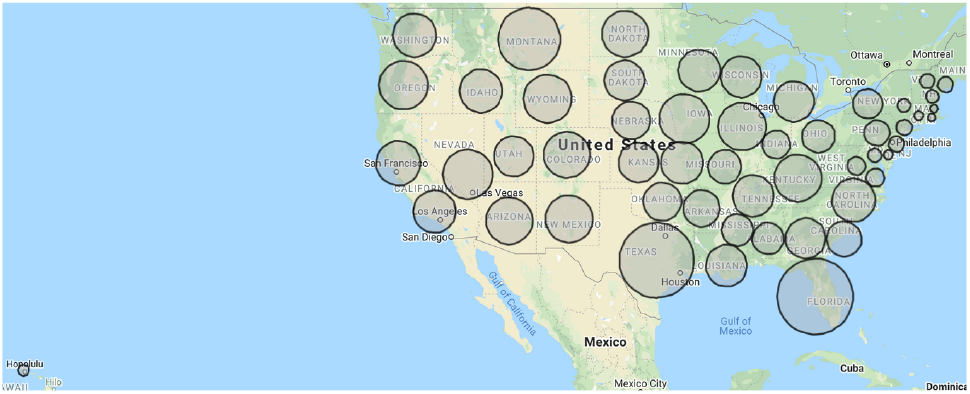
States of USA

### C. Preprocessing Data

- Uppercase to lowercase
- Eliminate alphanumeric symbols
- Eliminate words with size less or equal than 3
- Add some exceptions

### D. Visualization

- Number of retweets and kind of device (Android or iOS)
- Hour distribution per day and during the week
- Genre distribution and tweets per day by Genre
- Cloud of words and bigrams by Genre
- Tweets per day and during the week
- Histograms with the top 30 of the most posted words per day and during a time period
- Cloud of words to analyze the most frequent terms per day of the technical conferences held by the Mexican government

## III. Results

Due to the spread of the disease in the world, social media platforms and news websites have become places where there is an intense and continuous exchange of information between government agencies, professionals and general public.

### A. Frequency of tweets per state

For the respective analysis, a review of the most relevant news regarding this outbreak of black death was made. Due to the pandemic stage that we live based on COVID-19, it had to be differentiated from tweets on black death, so it had to be filtered by keywords. In figure 2, we look at the number of tweets per state. While California is where the Black Death case occurred, it is New York where this is most often discussed, followed by Texas and Washington.

**Fig. 2.**
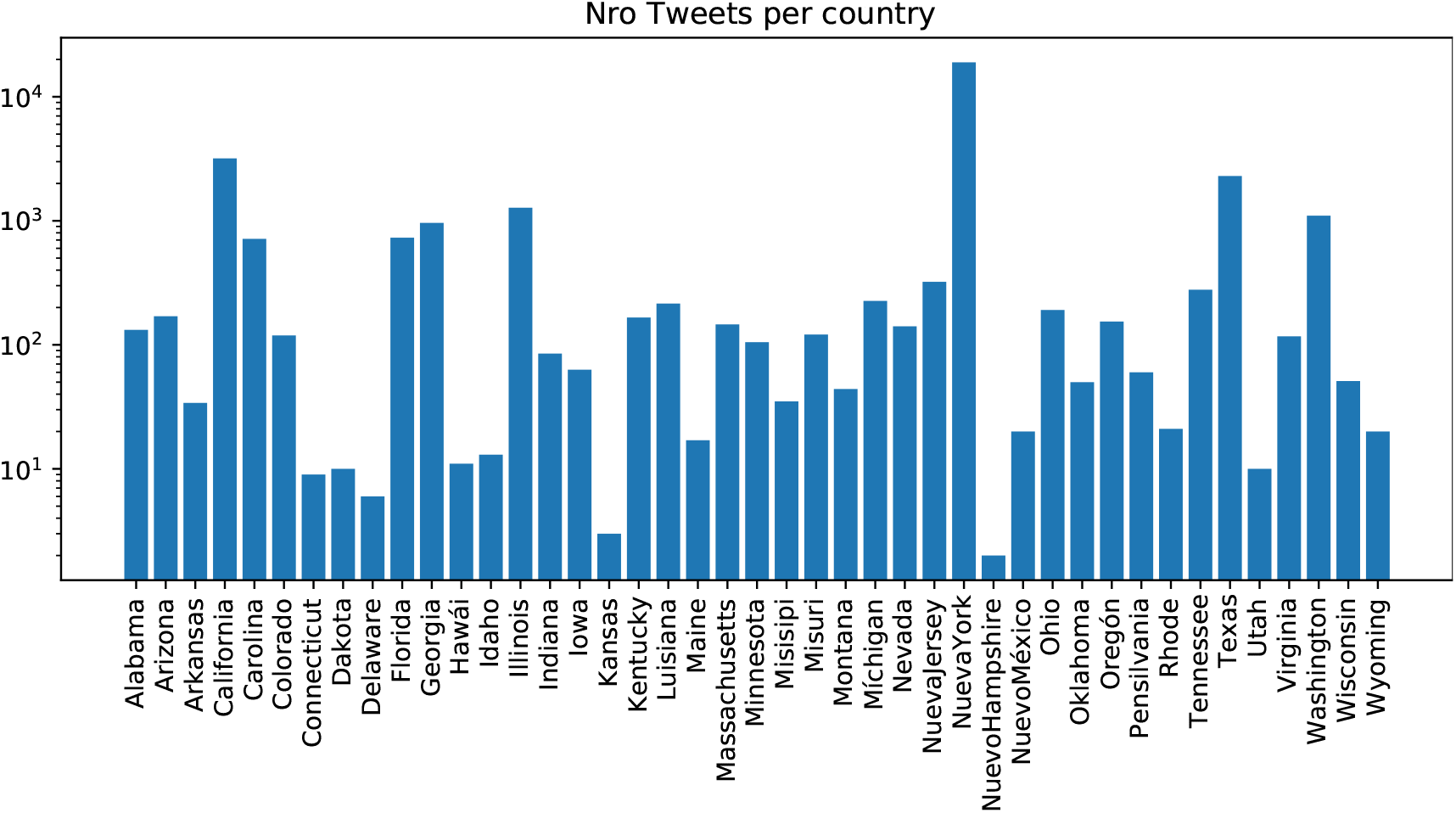
States of USA

It is noted that the interest of the population, based on the number of tweets, is in the cities with the highest level of cases of COVID 19. According to Google statistics [9], the cities that present the greatest number of cases are those that talk the most about this issue. This indicates that there is a convergence of interests on the conjuncture, official news and users of social networks. Another important point for users from the cities most affected by COVID 19 to comment more on this news about Black Death, is that, in each of these states, there has been more information about this type of zoonotic diseases outbreak, which can lead to increased anxiety and stress caused by the pandemic, as well as by the efforts to reduce its spread [10].

### B. Frequent terms

Twitter is a two-way communication platform with a social network nature that limits its messages to 280 characters and, although it seems very limited. it is widely used for more formal topics than other social networks. In addition, it allows to generate trends based on the use of hashtags (#) which makes visible important issues that involve public health. A word cloud was generated, which is shown in Figure 3, which gives us a visualization of the 50 most frequently used words in the study time range. We observe that the topic of the origin of the Black Death is the greatest interest of people in all USA. In order to have more detail, the same procedure was performed for all states of the USA, which is shown in figure 4, where we observe that the word “Black Death” is present in most of them. These words related to death are linked to a fear generated at this stage by the COVID-19 pandemic, so it is a wake-up call to examine the psychological consequences of stressors arising from problems related to the emergence of new diseases [11].

**Fig. 3.**
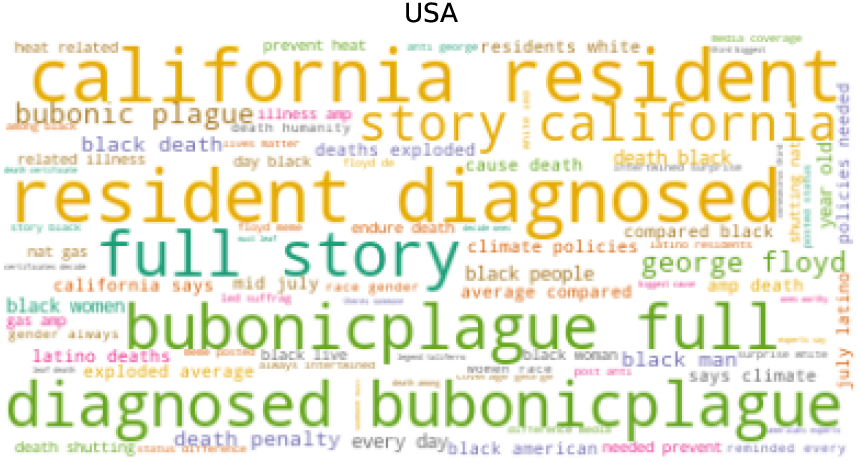
States of USA

**Fig. 4.**
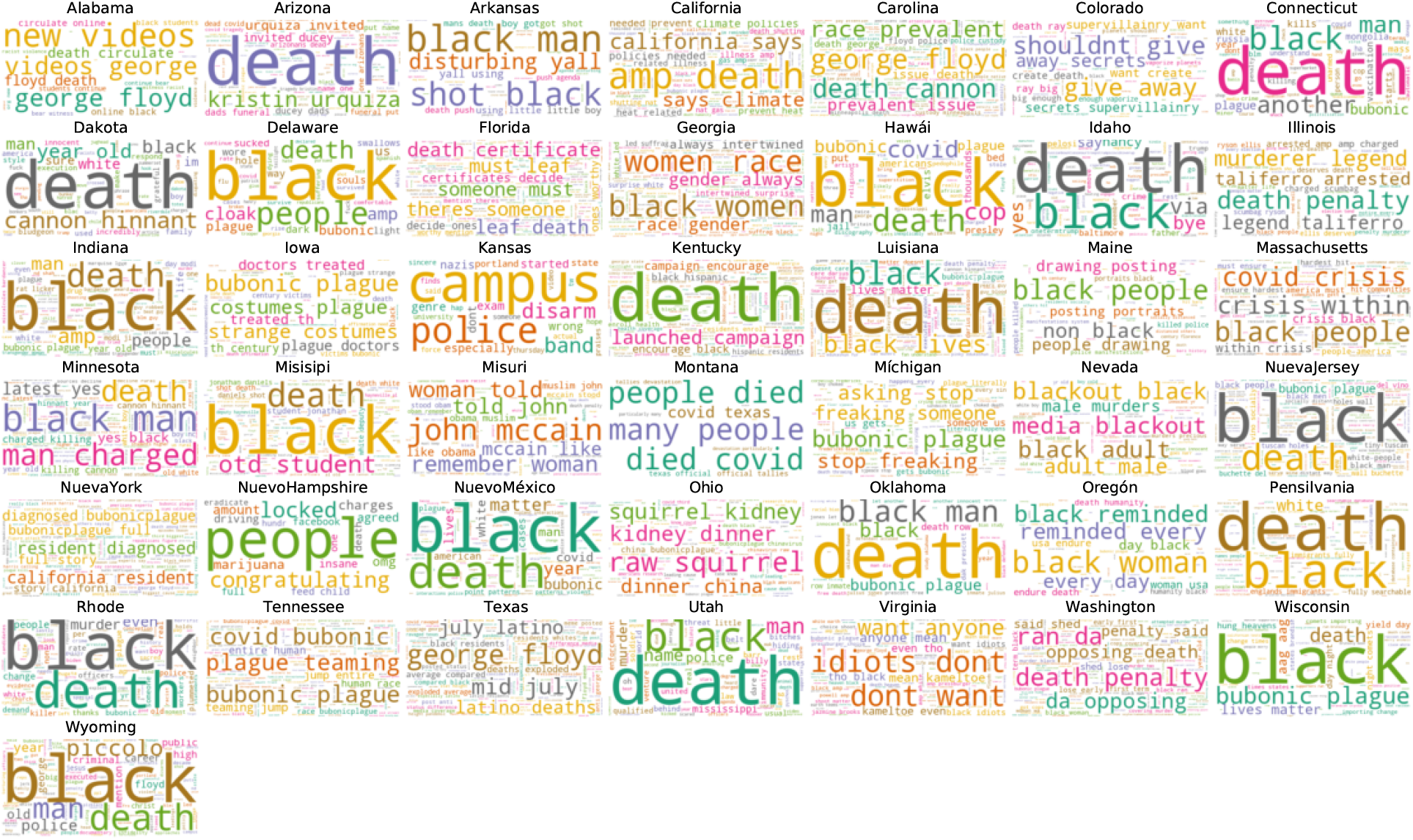
States of USA

Also, in the analysis of words, we can see that users talk about medical treatment, control procedures and political issues related to the health sector.

The heat map is a very useful visual form to know the variables and their relationships, so it is used in Fig. 5 as a matrix of the feeling of users through their tweets 24 hours a day. The values are averages and show there are states with greater permanence of the idea of the Black Death as a reality in their country, such as the case of California, Florida, Texas, and New York having the highest 24-hour rate.

**Fig. 5.**
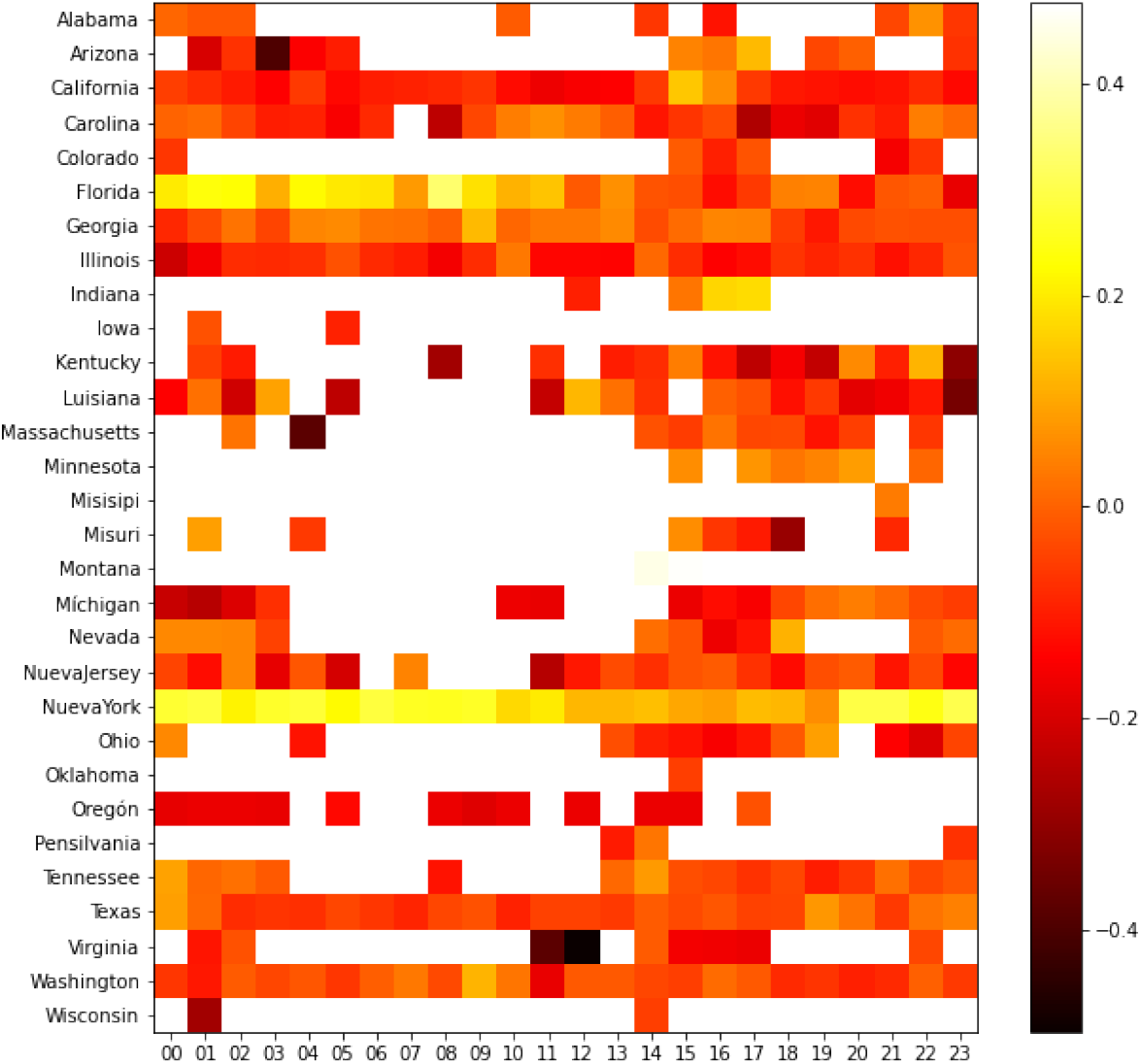
States of USA

It is also evident that the hours to create tweets, at the level of all states, is from 16 hours to 03 hours, being the period of greatest activity in this social network.

However, because the Internet is an interactive medium, there is also the potential to seamlessly collect even richer data from people, or to direct them to interventions, this represents nothing less than a paradigm shift, as traditional surveillance efforts, which are based, for example, on monitoring emergency room admissions or over-the-counter drug sales, happen without consumers even noticing it or being able to provide input. In contrast, using infoveillance methods, consumers can be directed to provide additional information [12].

In stages like these ones, where epidemiological surveillance is looking for new, timely and real-time detection alternatives, it is important to evaluate options, such as the use of social networks. Besides, the World Health Organization (WHO) has already stated that “the cornerstone of public health security aims to detect high rates of disease and mortality, the implementation of control measures and the notification to the WHO of any event that may constitute a public health emergency or international concern” [13]

Black Death, currently, does not have an effective vaccine and its mortality rates are 30 % to 50% when left untreated, while pneumonic plague, when left untreated, is always fatal. Together with this, the effect of Black Death worldwide may be undermeasured in data due to unrecognized cases and the fact that countries do not report them [14]

In this context, the ask of epidemiological surveillance is motivated by the notion of public health, so the analysis of alternatives, such as the use of Twitter for the rapid detection of trends related to this topic is important. In addition, it can be bidirectional as it could also be used to build public health campaigns involving specific Twitter users and with the specific information users require.

## IV. Conclusions

The use of Twitter has many potentialities and applications for determining the level of attention to critical situations, such as the presence of a Black Death case. Besides, it can be used as a data source in semi-real time and with a significant sample. This, promoted by the massive use of smart devices, so along with the development of machine learning tools can become a useful tool before, during and after events related to public health.

## V. Recommendations

- It is important to remember that the collection of data is the first step in this process of analysis, then a careful selection of keyword will impact the whole process, perform some experiment to find the proper terms.
- Preprocessing is important to clean data and set ready for posterior steps.

## Data Availability

The data will be released soon.

